# Biomarker discovery study design consistent with the Receiver-Operator Characteristic

**DOI:** 10.1101/2025.04.23.25326188

**Authors:** Joakim Ekström, Ivaylo Stoimenov, Jim Åkerrén Ögren, Tobias Sjöblom

## Abstract

The field of early biomarker discovery is characterized by a lack of consensus on the choice of statistical methodology, which may impede later progress towards clinically useful biomarkers. The Receiver-Operator Characteristic (ROC) is a de facto standard for determining the accuracy of *In Vitro* Diagnostic (IVD) devices. We demonstrate a biomarker discovery study design that achieves endpoint-consistency through use of ROC analysis from study objectives through sample procurement plan, sample size determination, to data analysis. Through simulations, the investigator can be informed on suitable study size to demonstrate an effect superior to the current best clinically used biomarker for the purpose. The study design is illustrated using proteomic data of newly diagnosed cancer cases and concurrent external controls, and statistically significant composite biomarkers are validated using independent data generated using the same proteomic analysis method. Intriguingly, commonly used feature selection methods do not identify the same composite biomarkers from the same data, and their selections show limited overlap with the ROC-based analysis. The proposed approach can facilitate translation of scientific discoveries into regulatory approved biomarker tests fit for use in clinical medicine.

## Introduction

Over the past four decades, significant advances in genomics, proteomics, and metabolomics have enabled precise measurement of a vast array of analytes, spurring hope for the discovery of new biomarkers for novel *in vitro* diagnostic (IVD) devices [1]. Since clinically useful biomarkers have been utilized for diagnosing various metabolic disorders, the focus has naturally shifted to more complex diseases, such as cancer [2]. However, despite substantial economic investments, clinically useful biomarkers for early detection are still lacking for the majority of cancers [3]. Efforts toward multi-cancer early detection have proven to be even more challenging [4]. Among the reasons discussed for biomarker study failures is the inappropriate selection of sample donors [5], small sample sizes [6] poor analytical method [7], inappropriate statistical analysis, inadequate handling of multiplicity in hypothesis testing [8], and overlapping training and validation cohorts [9]. Any flaw in the study design may ultimately lead to failure in obtaining relevant regulatory approval for clinical use and restrict the biomarker’s applications.

While the scientific literature lacks a universally accepted definition of the term ‘biomarker’, regulatory authorities provide working definitions that guide the clinical phase of biomarker development [10]. These definitions prioritize biomarker tests that correctly identify individuals with the condition (positive diagnosis) and those without it (negative diagnosis). The proportion of individuals with the condition who are correctly diagnosed is termed sensitivity, while the proportion of individuals without the condition who are correctly identified is termed specificity [11]. The sensitivity-specificity pair, known as the ROC point, serves as the primary measure of diagnostic accuracy. Collectively, these ROC points form the ROC curve, from which diagnostic performance can be summarized in a single metric: the ROC AUC (Area Under the Curve) [12]. Since demonstrating that a new biomarker test has non-inferior diagnostic accuracy compared to an already approved predicate device is crucial for regulatory approval, it is prudent to benchmark against such state-of-the-art devices early in the discovery phase. The ROC should be the statistic of choice, as it is widely used to assess the effectiveness of diagnostic tests [13].

In applications where individual biomarkers lack sufficient accuracy to address diagnostic challenges, combining multiple biomarkers offers a potential solution [14]. Several methods of combining biomarkers exist, however there is unsatisfactory cohesion in the use of terminology used to refer to the various methods. This issue was highlighted during a recent public meeting organized by the FDA aimed at developing standardized terminology for composite biomarkers [15]. Further, there is a need for stringent approaches to study dimensioning and design that enable informed decision making.

Here, we propose and demonstrate a biomarker study design guided by regulatory standards and best practices, aiming to circumvent common pitfalls highlighted in the biomarker literature [6]. Using assumptions based on the performance of approved IVD devices available at the time of writing, we conduct statistical power estimation via Monte Carlo simulation [16]. This approach provides insights into optimal sample sizes, data allocation between training and validation cohorts, and the number of candidate biomarkers to advance from the training to validation phase.

## Results

### Principal steps for adequate and well-controlled studies

There are several benefits in designing a biomarker study so that it incorporates many of the characteristics of a clinical trial of regulatory standards, which are codified in, e.g., Title 21 of the U.S. Code of Federal Regulations (CFR) part 314.126 [17], explained in International Council for Harmonization of Technical Requirements for Pharmaceuticals in Human Use (ICH) [18] and incorporated in U.S. Food and Drug Administration (FDA) guidance documents [10].

From these relevant regulatory documents, a workflow for design of biomarker discovery studies can be derived (Fig. 1). Title 21 CFR (314.126) proposes that the objectives of the study are articulated clearly in the early process of study design (Fig. 1A) [19]. A clearly stated study objective will promote coherent decision-making throughout the study design process. Examples of biomarker study objectives are to validate a biomarker that predicts complete remission of colon cancer subsequent to local resection or explore biomarkers that detect ovarian cancer in vaguely symptomatic patients. It is advisable to designate primary objectives and possibly secondary objectives, and check that objectives are not mutually contradictory, e.g. an objective that is simultaneously both confirmatory and exploratory.

**Fig. 1.**
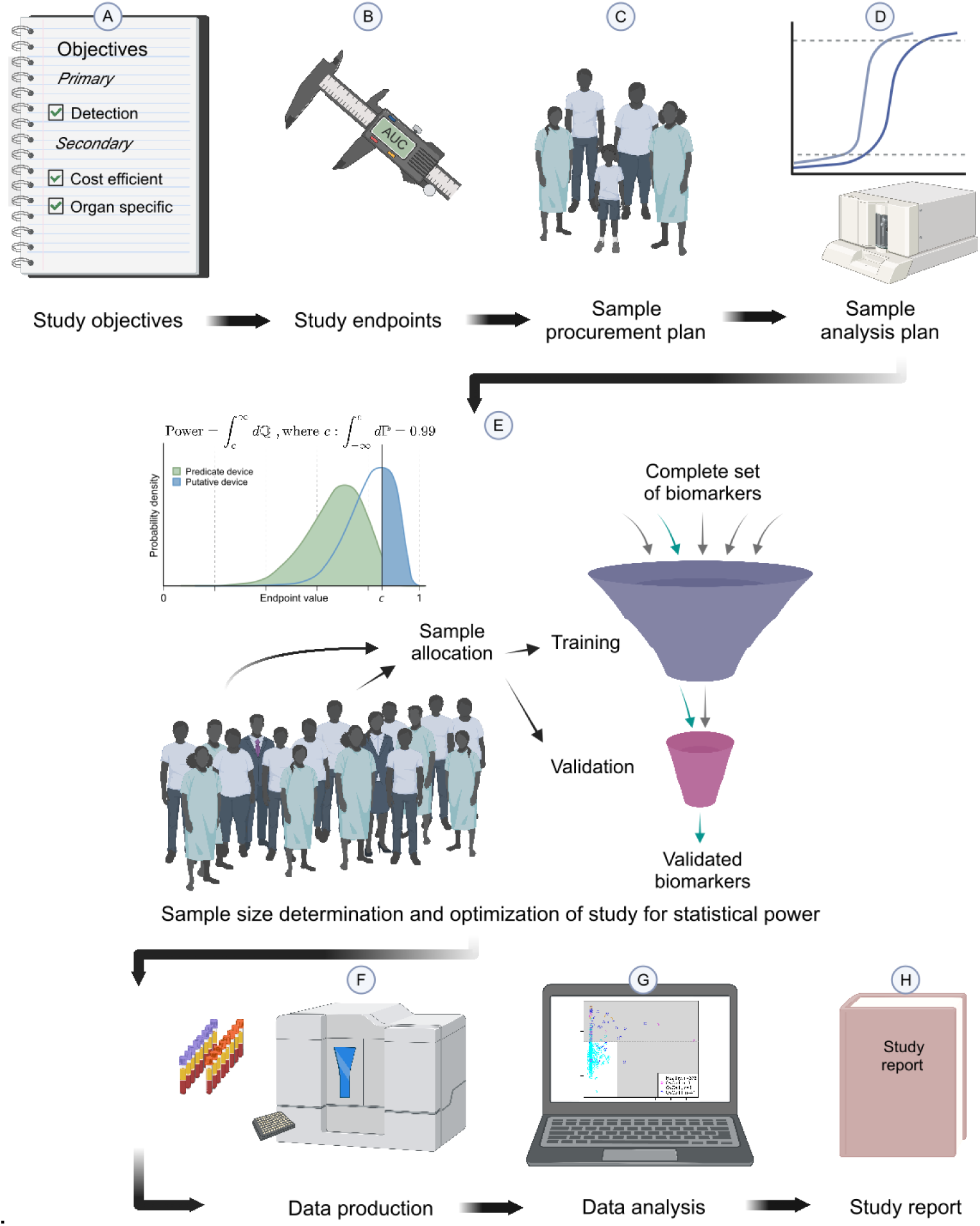
Steps of a biomarker discovery study. An example protocol for an adequate and well-controlled study, partitioned in key steps for a visual clarity. **A.** First, the primary and secondary study objectives are clearly defined to enable sound and cohesive decision-making. **B.** The endpoints corresponding to each of the study objectives, such as superior sensitivity, specificity and ROC AUC to the state-of-the art biomarker for early detection of a specific cancer type in blood drawn at diagnosis are defined. **C.** Next, a plan to procure samples from a sufficient number of cases and controls relevant to the study objectives and the intended use, is prepared. **D**. Selection of technologies for sample analysis with clinical translation in focus, while considering factors such as dynamic range, CV, reproducibility and sample scaling. **E.** Computation of statistical power to determine optimal study size and the sample allocation to discovery and validation steps given the study objectives. **F.** Once the sample size is known, relevant samples are procured from biobanks or collected *de novo,* and data is then produced according to best practices introducing minimal bias. **G.** The data are analyzed for primary and secondary study endpoints, for example by computing the ROC AUC of all combinations of single and composite biomarkers and performing hypothesis testing for superiority versus a state-of-the-art biomarker. **H.** The study report is prepared according to regulatory guidelines.

Next, endpoints are defined toward the evaluation of the performance of the biomarker, usually corresponding to the study objectives. Typically, the efficacy endpoint of a biomarker is its diagnostic accuracy, and the Receiver-Operator Characteristic (ROC) Area Under curve (AUC) has become a de facto standard statistical measure for this purpose, see e.g. FDA Guidance [10], although in some instances other endpoints could be more relevant, such as a particular sensitivity and specificity pair (Fig. 1B). It is practical to test statistically for superiority of the observed biomarker AUC-value relative to absence of diagnostic ability, i.e. AUC-value of 0.5, or superiority relative to the AUC-value of an appropriate predicate device [10].

Procurement of samples is typically one of the most resource-intensive parts of a biomarker study (Fig. 1C), and several pitfalls discussed in the literature pertains to sample procurement [14]. Adequate assurance that cases have the condition studied, e.g. cancer, and that controls lack the condition, is crucial [17]. To this end, disease registries or electronic health records serve as a valuable resource for guiding decisions on sample inclusion [20]. Identification of subpopulations with relevant pathologies can also contribute to the study outcomes [14]. Inappropriate or inadequate selection of controls is a known pitfall [19, 21], also discussed in 21 CFR [17]. Importantly, the purpose of a controlled study is to distinguish the weight of the desired effect from common influences such as bias. Biases and potential confounders such as age, sex, administration of drugs or therapies, and diseases other than the condition studied are a particular challenge in externally controlled designs [14, 17]. Uniformity in sample handling and storage is discussed by Drucker and Pavlou, and a proposed solution is to predefine SOPs to minimize preanalytical biases [14, 19]. Blinding is a suggested measure to further minimize bias [17].

Appropriate analysis of the samples is thoroughly discussed in the literature, and is potentially a contributor to problems with reproducibility [22] (Fig. 1D). Analyses which are difficult to reproduce due to high coefficient of variation, measurements of relative nature, or other reproducibility challenges will often limit the value of biomarker study since validation then is intrinsically challenging. It is recommended to use reliable and trusted analytical techniques [22].

Sample size determination and statistical power optimization are essential steps conducted prior to data acquisition (Fig. 1E). Based on the chosen analytical methods, samples may be divided into training and validation sets or used in their entirety for biomarker discovery. A common approach involves allocating a larger proportion of samples for training, while using a smaller set to validate hypotheses. However, certain analytical methods achieve optimal biomarker discovery by incorporating all available data [16]. Accurately estimating statistical power is a critical step before initiating sample collection and data acquisition. If the desired outcomes cannot be achieved with the available sample size, additional samples must be secured, or the study goals should be adjusted to reflect such limitations.

The quality of input data is of profound significance for the study outcomes (Fig. 1F). Ensuring the purity of biological samples, with minimal contamination, is important; however, using controls, which do not have the disease, is even more critical. While technical replicates can partially account for the biological complexity of the samples, prioritizing a greater number of independent, unique sample donors is a more effective strategy. Selecting methods with a high dynamic range and enhanced precision, particularly for multiplex measurements, often yields superior results [23].

After data acquisition, the use of appropriate analysis tools is crucial (Fig. 1G) [22]. From the perspective of health regulators, deterministic analytical tools are generally preferred over highly dynamic and less predictable methods, such as those involving machine learning and artificial intelligence, since certain aspects of the AI/ML tools are met with skepticism [24]. The use of ROC analysis is a well-established methodology that complements filing documentation for biomarker evaluation. It provides a clear framework for comparing the direct performance of a prospective biomarker, including the true positive rate and the associated penalty of the false positive rate, against any predicate device at various thresholds.

The choice of statistical method is a documented pitfall discussed by Pavlou and Drucker, also covered by 21 CFR [14, 17, 19]. Evaluating the true statistical significance of prospective biomarker discoveries is crucial for their validation and clinical performance. However, stringent statistical criteria, while ensuring robustness, can also hinder the progression of promising biomarkers to subsequent stages, potentially reducing validation failures but limiting innovation [7]. Balancing rigor with practical flexibility is therefore essential in the biomarker discovery process.

The analysis data and study outcomes should be shared with the scientific community in a format that allows for thorough evaluation of the results and interpretation of the biomarker’s applicability within a broader context (Fig. 1H). Findings should be presented alongside a significance estimate, clearly specifying the significance level used.

### Dimensioning studies for discovery of composite biomarkers

In this work, we use the term unitary biomarker to refer to biomarkers, i.e. characteristics in the body or bodily products that can be measured objectively, that are constituted by a single measurement, and composite biomarker to refer to a biomarker that is constituted of two or more measurements. We use the phrase constituent biomarkers to refer to the unitary biomarkers that constitute a composite biomarker. The subcategories “integrative composite biomarker” and “classification composite biomarker”, which refer to composite biomarkers that are applied, respectively, by dimension reduction to a single value on which a cut-off is applied, and by maintaining dimensions and applying cut-offs to the multi-dimensional observation of the composite biomarker, have been proposed [15].

We have used the method of Prakash and implemented by Banyan Brain Trauma Indicator (DEN170045, also K201778) using one cut-off per constituent biomarker, and extended it to combinations of arbitrary numbers of biomarkers (see Methods) [25, 26]. Further, we implemented the composite biomarker method such that positive classification is permitted when any constituent is above its cutoff, like the Banyan BTI, and when all constituents are above their cutoffs, like Prakash (Suppl. Fig. 1). Also, positive classification is permitted when levels of the constituent biomarkers are either elevated or depressed. The P-values relative to superiority of observed AUC-values vis-à-vis a predicate device were determined using exact computation, and correction for testing of multiple hypotheses was done using the Bonferroni method [16].

### Optimization of study design

The pitfall of small sample size is discussed by Yotsukura, Drucker and Pavlou [14, 19, 21]. Based on regulatory approval procedures, it is necessary to have substantially equivalent ROC performance to existing tests for the same purpose in order to introduce a new biomarker test. As clinical use of a new biomarker test likely requires better performance than the state-of-the-art biomarker, we here simulated the discovery of biomarkers with significantly better performance than an existing biomarker with regulatory approval. Epi proColon (ROC AUC 0.82) is a blood biomarker for early detection of colorectal cancer with FDA pre-market approval [27]. The FDA recently approved Guardant Shield, a blood test for detecting advanced stages of colorectal cancer, demonstrating 83% sensitivity and 90% specificity for advanced neoplasia thus approaching the performance of the stool-based test ColoGuard (ROC AUC of 0.93) [28] (Fig. 2A). However, Guardant Shield has not yet been adopted into routine clinical practice [29, 30].

**Fig. 2.**
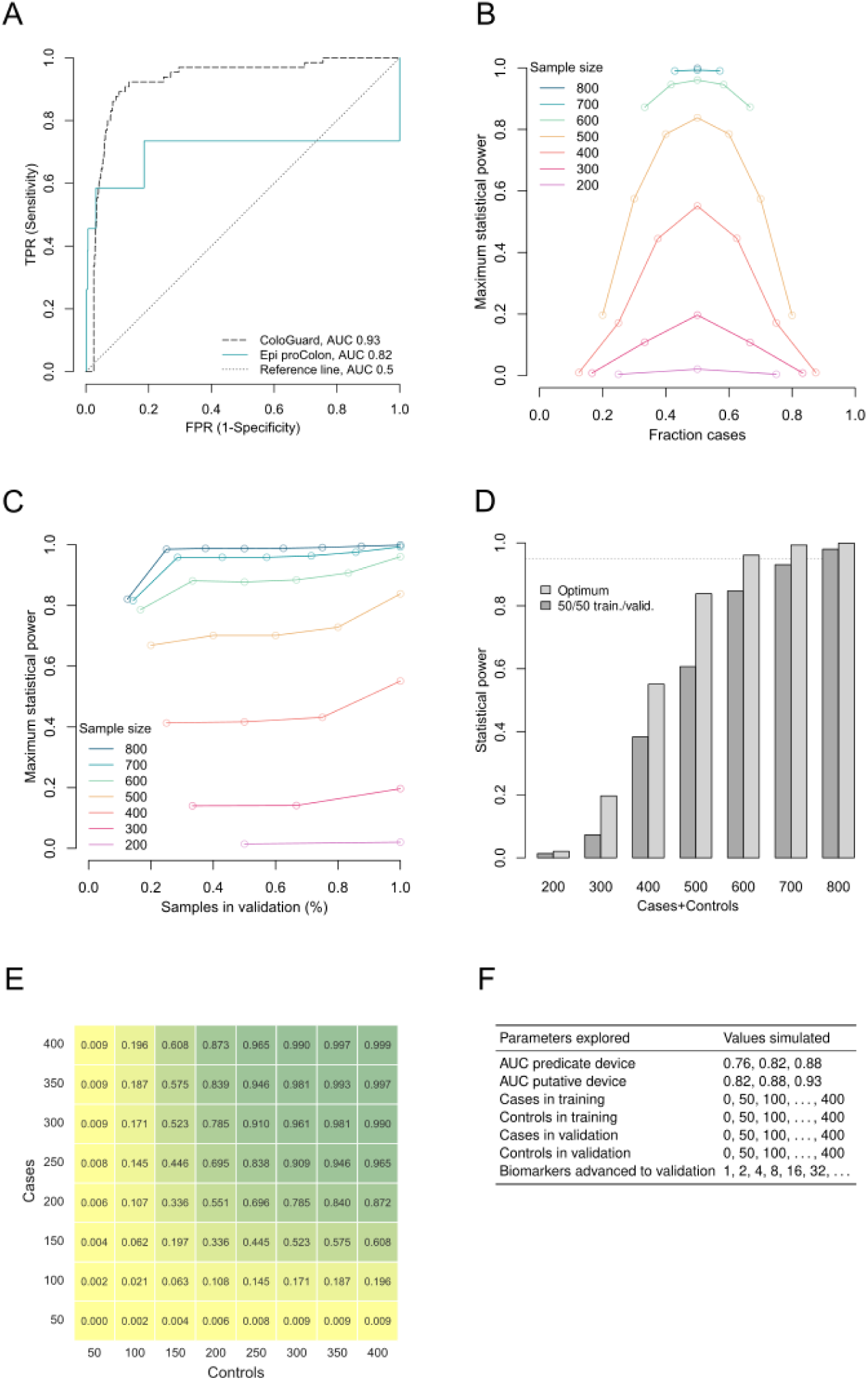
Determining optimal study design and sample size for biomarker discovery studies using statistical power simulation of the ROC AUC. Monte Carlo simulation of composite biomarker discovery in data encompassing 165 proteins in combinations of 4 and the primary endpoint to demonstrate superiority of a putative biomarker to a state-of-the-art biomarker. **A.** Superiority of a single putative biomarker with performance on par with ColoGuard (ROC AUC 0.93) relative to a predicate device biomarker on par with Epi proColon (ROC AUC 0.82) at 99% statistical significance is simulated. **B.** Statistical power simulation of the impact of the fraction of cases of all samples in the study. **C.** Statistical power simulation under a range of fractions of samples allocated to validation in a training/validation study design. **D.** Statistical power simulation of allocation of the same number of cases and controls either to a conventional training/validation study design (50% training, 50% validation) or an optimized design (100% validation). **E.** Statistical power as a function of the fraction of cases and controls in a study design with 100% of samples allocated to validation. A study size of 300 cases and 300 controls provides >95% statistical power. **F.** Parameters and values explored in simulations.

To find the appropriate study design and size, we conducted Monte Carlo simulations of unitary biomarkers in combinations of up to 4, across a range of sample sizes allocated to training and validation sets (see Methods). The desired statistical power was set to 95%, when using the 99% statistical significance level. As expected, increased sample size led to more sharply defined AUC-value probability distributions and improved statistical strength (Fig. 2B and Suppl. Fig. 2).

In all scenarios simulated, the greatest statistical power was obtained when all data were allocated to the validation set (Fig. 2C). Further, the statistical power was optimal for a given total number of cases and controls when the number of cases and controls were equal (Fig. 2B). Compared to a design of a training and validation set with half of cases assigned to each, allocation of all cases to a single validation set markedly improved statistical power (Fig. 2D). For this example, 300 cases and 300 controls provided statistical strength >0.95 for finding a true effect (Fig. 2E). These findings were reproduced under a range of assumed putative and predicate biomarker AUC-values (Fig. 2F). Although the number of classification composite biomarkers was large, as determined by the binomial coefficient *n* choose *k*, it remained finite. This allows for the correction of multiple hypothesis testing across the full range, from a single composite biomarker to the entire set. Taken all together, using an optimized study design, satisfactory statistical power can be obtained at reasonable sample sizes, and by conducting the statistical power estimation, the known pitfall of small sample sizes can be mitigated.

### Impact on statistical power of using separate versus pooled controls

When samples and data are collected to develop biomarkers for several different conditions or tumor types, there may be instances where several groups of non-diseased controls are generated within the same study [31]. It is valuable to understand how statistical power is affected if these control groups are pooled. As expected, the statistical power will always increase with the addition of more controls, albeit with diminishing returns (Fig. 3A). Pooling control groups also increased the statistical power to near the effect of having dedicated controls and is often preferred from a practical point of view (Fig. 3B). Pooling of a few large control groups incurred a lower such penalty than pooling several small control groups. If controls are pooled and re-used for several tumor types, the multiple hypothesis correction will reduce statistical strength beyond an optimum total number of controls.

**Fig. 3.**
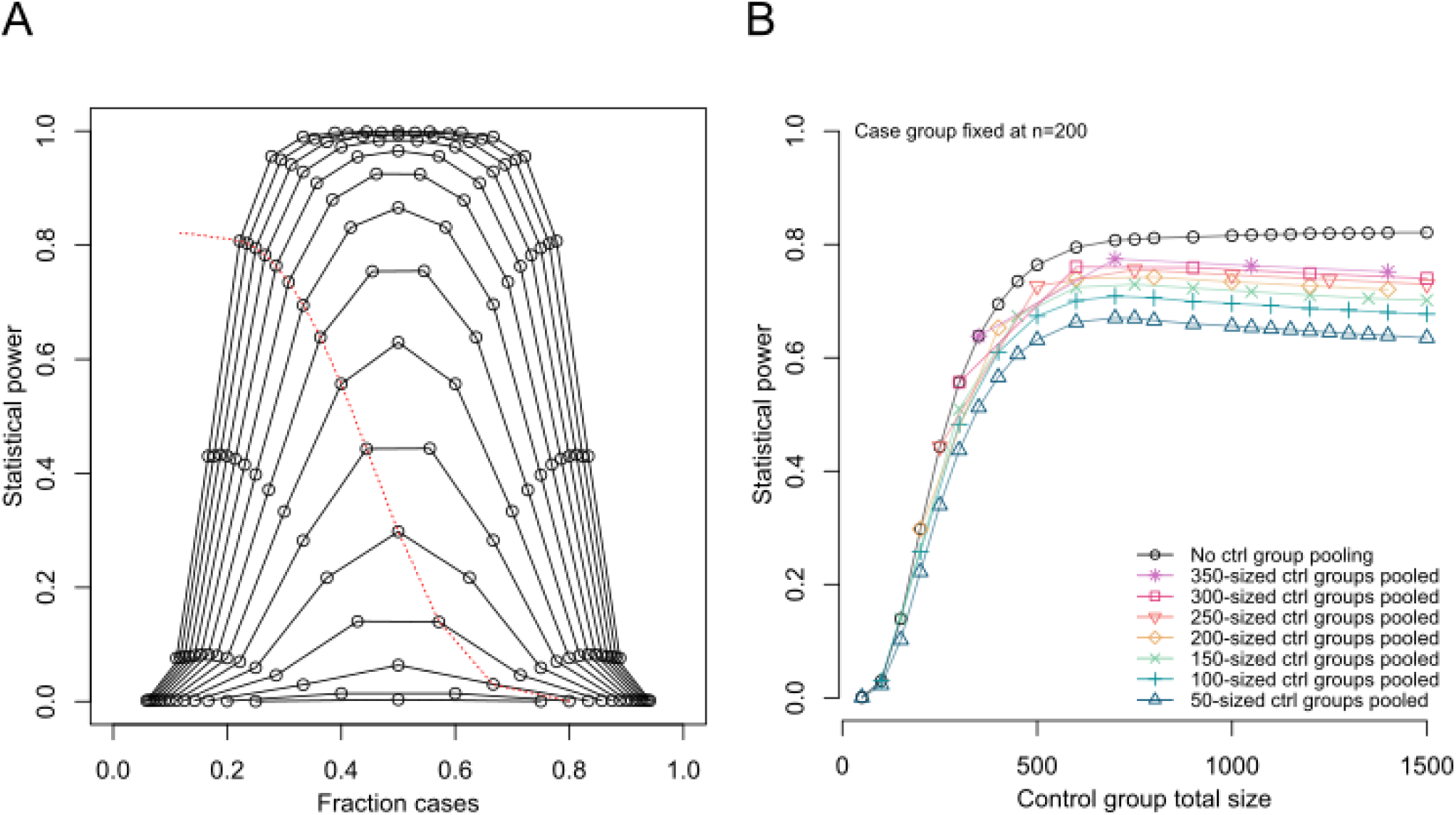
Increased statistical power from addition or informed pooling of controls. In a study where several different tumor types with corresponding matched controls have been collected under similar conditions, they can either be analyzed by tumor type versus its matched controls or versus all controls. Superiority of a single putative biomarker with performance on par with ColoGuard (ROC AUC 0.93) relative to a predicative device biomarker on par with Epi proColon (ROC AUC 0.82) at 99% statistical significance is simulated in a study of 165 proteins in combinations of 4. **A.** Addition of controls improves statistical power. Black lines, total number of cases and controls from 200 to 900 in increments of 50. Red dotted line, 200 cases. **B.** Pooling of controls increases statistical power, but with a penalty incurred for multiple hypothesis correction. Case group fixed at 200 with non-pooled controls (black line, corresponding to red dotted line in A) or controls pooled from groups of the indicated number of controls.

### Application for biomarker discovery

Biomarker development is illustrated through proteomic biomarkers for early detection of cancer. The publicly available data set from the CancerSeek study was used to develop biomarkers for early detection of cancer [31]. The dataset includes measurements of 39 proteins from 1, 004 individuals newly diagnosed with cancer and 812 healthy controls. Cancer diagnoses cover breast, colorectal, esophageal, liver, lung, ovarian, pancreatic, and stomach cancers. Additionally, a ninth, pan-cancer category is created by combining these eight cancer types. First, the statistical power to discover one effect on par with ColoGuard in a background of Epi proColon was simulated in each of the different diagnoses (Fig. 4). Given the number of controls, the sizes of the breast and colorectal groups were sufficient to achieve >95% statistical power (Fig. 4A).

**Fig. 4.**
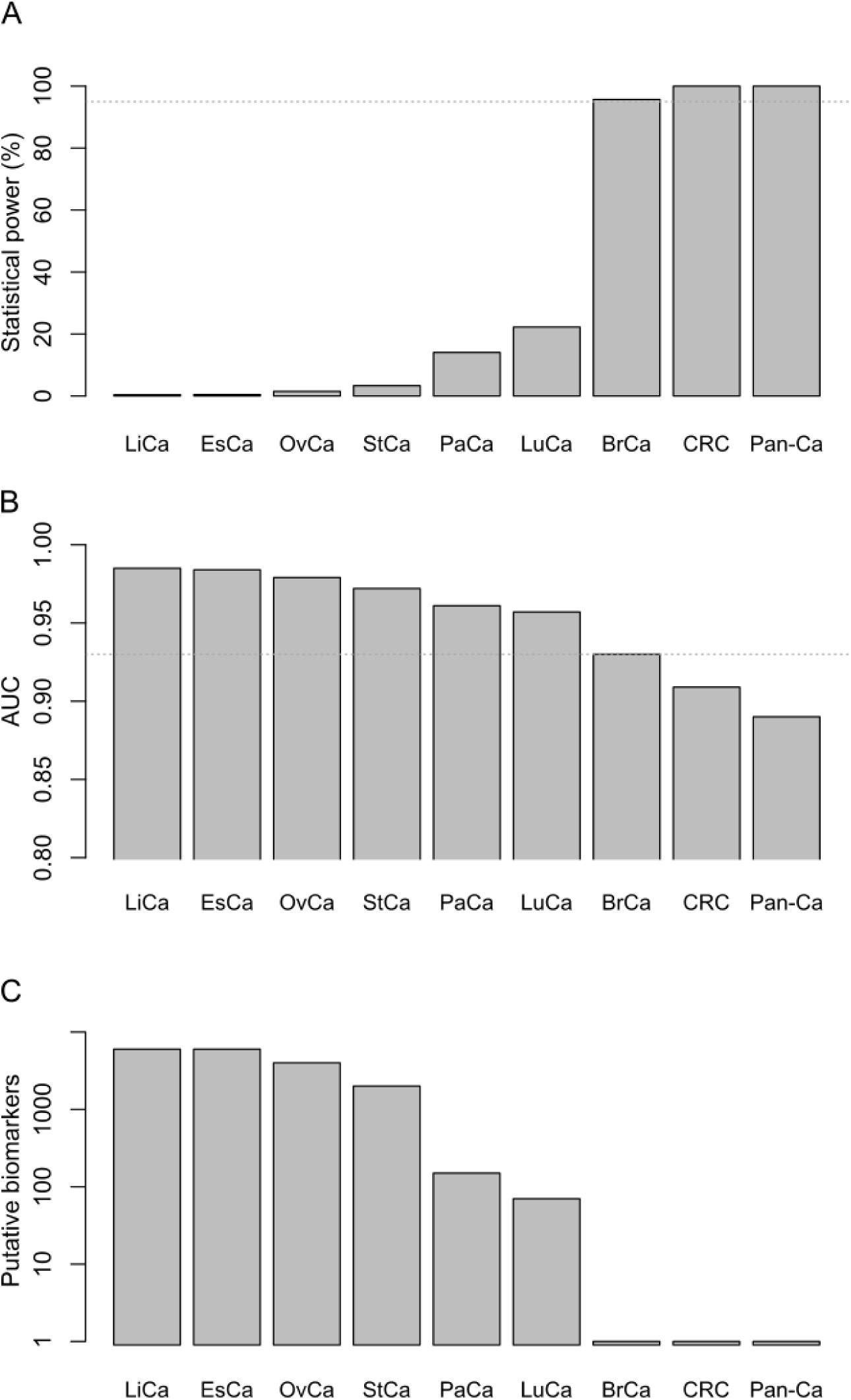
Statistical power estimation for CancerSeek cohorts. The statistical power was simulated in the different tumor type cohorts (range 44-388 cases) and all cases (PanCa, 1004 cases) of the CancerSeek study using all controls (800) pooled. **A.** Monte Carlo simulation of statistical power to find a single biomarker on par with ColoGuard (ROC AUC 0.93) relative to a predicate device biomarker on par with Epi proColon (ROC AUC 0.82) in data encompassing 39 proteins in combinations of 4. **B.** The ROC AUC value of a putative biomarker needed to achieve 95% statistical power by tumor type. Dotted line, ROC AUC of ColoGuard (0.93). **C.** The number of putative biomarkers on par with ColoGuard needed to achieve 95% statistical power of detecting at least one of them. LiCa, Liver Cancer; EsCa, Esophageal Cancer, OvCa, Ovarian Cancer; StCa, Stomach Cancer; PaCa, Pancreatic Cancer; LuCa, Lung Cancer; BrCa, Breast Cancer; CRC, Colorectal Cancer; Pan-Ca, Pan-cancer.

Next, we asked what the ROC AUC of a biomarker must be to ensure that it is detected given the size of each tumor type cohort. To achieve 95% statistical power, a biomarker had to have ROC AUC > 0.95 in all tumor types except breast and colorectal cancer (Fig. 4B). As the breast and colorectal cancer sub-cohorts of CancerSeek provide sufficient statistical strength to detect a single biomarker on par with ColoGuard in a background of biomarkers on par with Epi proColon, they are adequately powered following the logic developed here (Fig. 4A). A single ColoGuard-class biomarker could be discovered in the breast or colorectal cancer groups with 95% probability, whereas tens to thousands of such biomarkers were required to achieve the same power in the other tumor types (Fig. 4C).

Next, we asked whether composite biomarkers with ROC performance significantly superior to a state-of-the-art predicate device for cancer detection in blood could be found in the CancerSeek data. Based on the design optimization (Fig. 2), all cases and controls were allocated to a single validation set, as this was found to be optimal in all simulated scenarios. Composite biomarkers were created by combining four proteins, using the single-cutoff-per-measurand method, resulting in a total of 23, 688, 288 composite biomarkers. The ROC AUC was computed for all composite biomarkers and tested statistically for superiority vis-à-vis Epi proColon (ROC AUC 0.82). The critical values, under Bonferroni correction for all 23, 688, 288 composite biomarkers were computed and composite biomarkers with ROC AUC surpassing this value were identified (Table 1). Of the 9 tumor type cohorts, 7 had biomarkers that were, with statistical significance, superior relative to a biomarker with performance on par with Epi proColon. From these, several tumor-type specific and pan-cancer biomarkers performed well also in lower stage tumors (Fig. 5). Using a Swedish data set for colorectal, lung, and ovary cancer (Åkerrén Ögren et al, submitted), produced by the present authors using identical Luminex xMap technology and reagents from the same supplier (Merck Millipore, St Louis, MI, U.S.), we performed an additional statistical significance test of the composite biomarkers significantly superior vis-à-vis Epi proColon in the CancerSeek data set. Because of incomplete overlap of proteins, a subset of the significant biomarkers was reproducible in the Swedish data set (Table 1). Of the colorectal cancer biomarkers, 3 (4%) were superior relative to Epi proColon, with statistical significance under Bonferroni correction, in both data sets. Of the lung and ovary cancer biomarkers, 6 (3%) and 7210 (58%) were superior relative to Epi proColon in both data sets. ROC plots for top performing biomarkers are shown in Fig. 6. Thus, the ROC-based approach to composite biomarker discovery identified biomarkers that could be reproduced also by external validation.

**Fig. 5.**
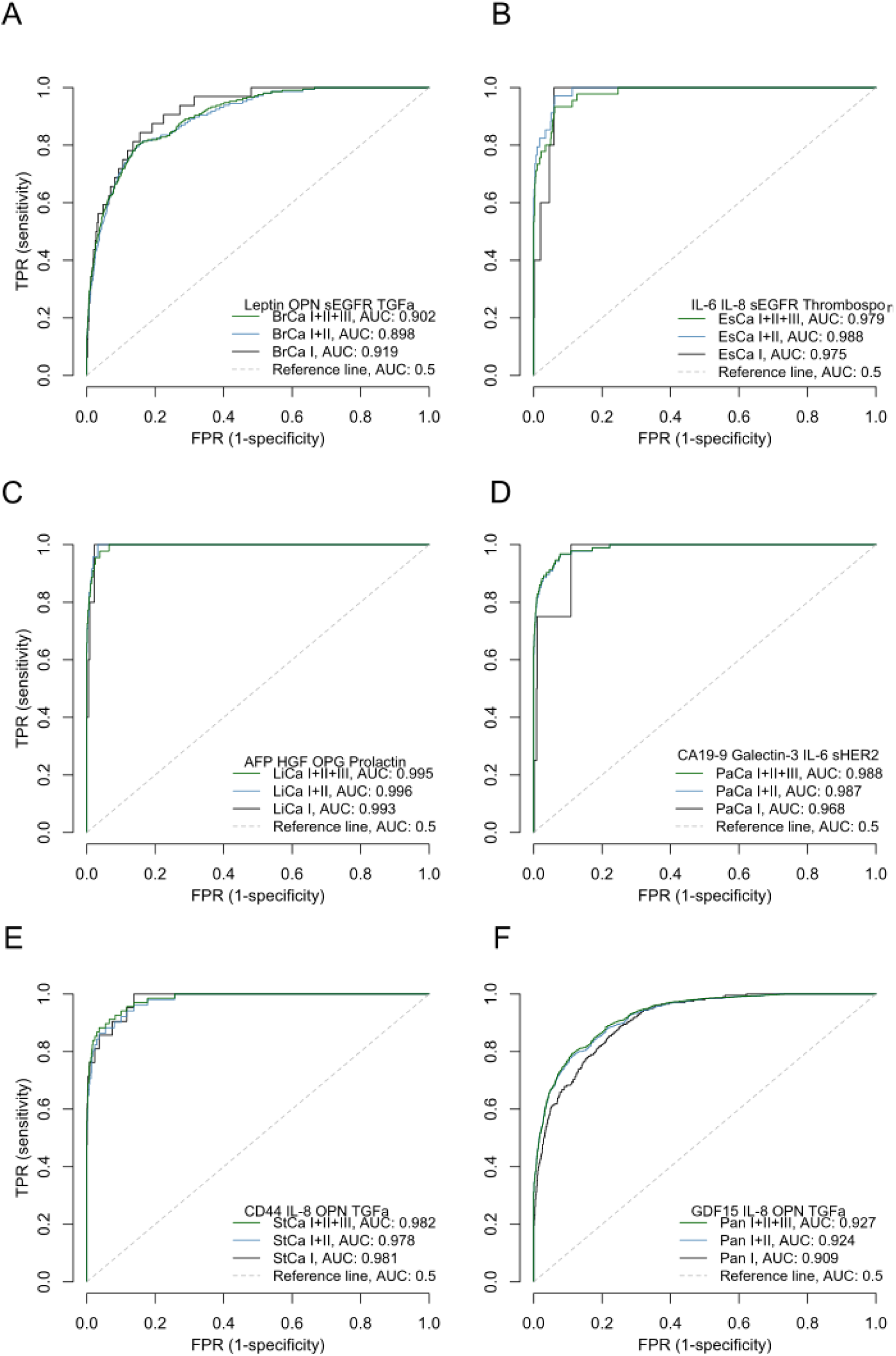
Four-protein diagnostic biomarkers from CancerSeek. The ROC AUC for all combinations of 4 proteins from 39 measured was computed and tested for superiority to Epi proColon (ROC AUC 0.82) using the proposed ROC-based discovery approach. The best performing biomarker as determined by ROC AUC for stage I-III tumors is shown with diagnostic performance by tumor stage for breast (BrCa) (**A**), esophageal (EsCa) (**B**), liver (**C**), pancreatic (PaCa) (**D**), stomach (StCa) (**E**), and pan-cancer (Pan) (**F**). The biomarkers in **C**-**F** had significantly better performance than Epi proColon (*p* < 0.01).

**Fig. 6.**
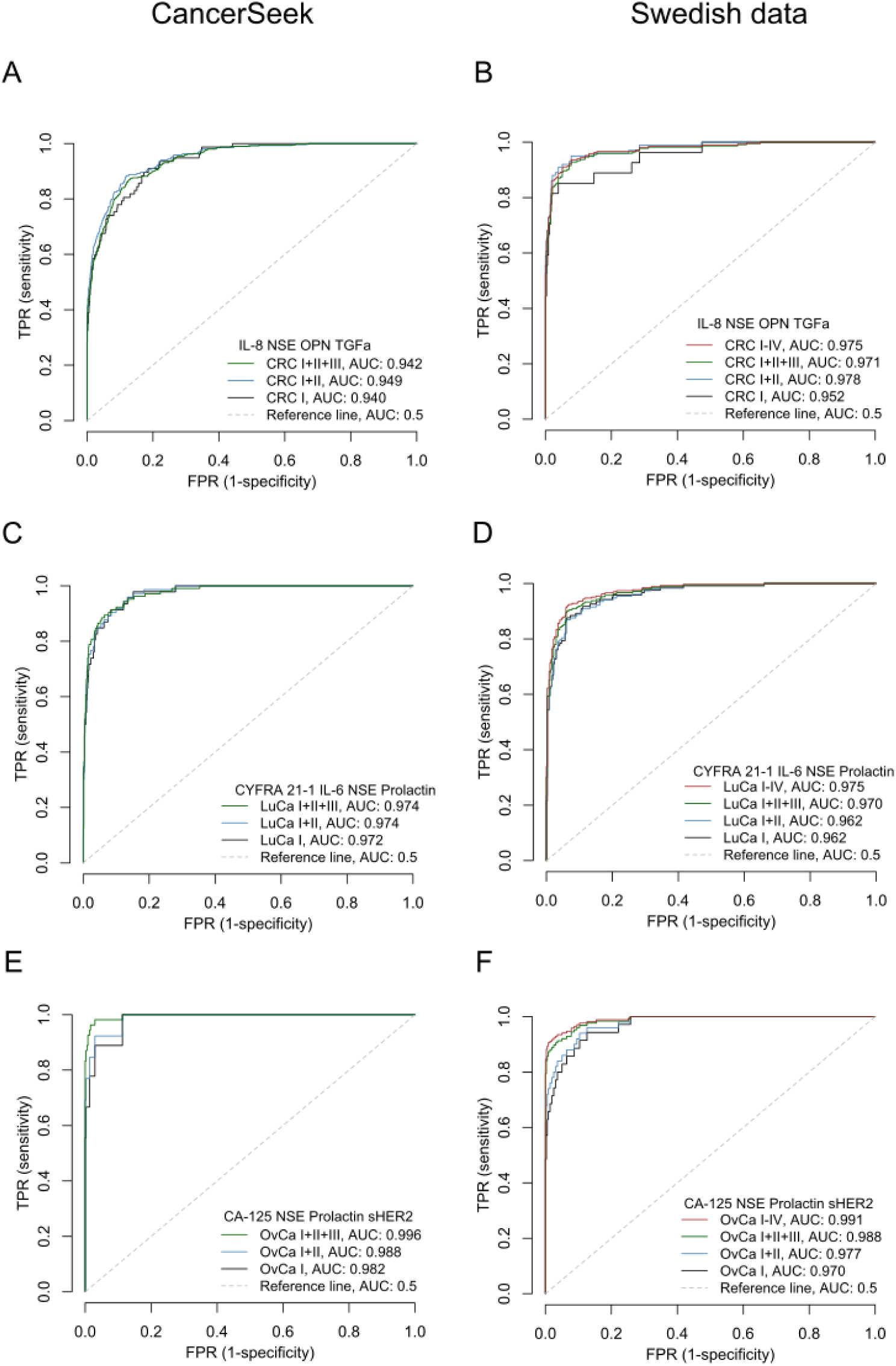
External validation of biomarkers for detection of lung, ovarian and colorectal cancers. The ROC of biomarkers significantly superior in performance to Epi proColon identified in CancerSeek (left panels) was determined in an external Swedish dataset (right panels) for colorectal (CRC) (**A** and **B**), lung (LuCa) (**C** and **D**), and ovarian cancer (OvCa) (**E** and **F**).

**Table 1.** Discovery and external validation of four-protein blood biomarkers for detection of cancer. The data set of CancerSeek, encompassing 39 proteins measured across 1004 individuals newly diagnosed with cancer and 812 healthy controls, was used to discover biomarkers for early detection of cancer. For ovarian cancer, only female healthy controls were included. Biomarkers of four proteins were formed using the single cutoff per measurand method of forming composite biomarkers, yielding a total of (39 choose 4) * 9 * 32 = 23, 688, 288 composite biomarkers. All composite biomarkers were tested statistically for superiority vis-à-vis Epi proColon, AUC 0.82. The critical values, under Bonferroni correction for all composite biomarkers evaluated in CancerSeek, are indicated. A subset of the proteins measured in CancerSeek was also determined in an external cohort of cases and controls, and was termed reproducible biomarkers. Critical values for superiority to Epi proColon were computed with correction for the number of reproducible biomarkers.

| Tumor cohort | CancerSeek |  |  |  | Swedish data set |  |  |  |  |
| --- | --- | --- | --- | --- | --- | --- | --- | --- | --- |
|  | Cases | Ctrls | Crit. value, 99% corr. | Number of significant biomarkers | Cases | Ctrls | Reprod. biom. | Crit. value, 99% corr. | Number of significant biomarkers |
| Breast | 209 | 812 | 0.952 | 0 |  |  |  |  |  |
| Colorectal | 388 | 812 | 0.939 | 75 | 268 | 697 | 72 | 0.880 | 3 |
| Esophagus | 45 | 812 | 0.986 | 0 |  |  |  |  |  |
| Liver | 44 | 812 | 0.986 | 1807 |  |  |  |  |  |
| Lung | 104 | 812 | 0.969 | 210 | 265 | 697 | 168 | 0.879 | 6 |
| Ovary | 54 | 372 | 0.984 | 12376 | 167 | 353 | 8780 | 0.907 | 7210 |
| Pancreas | 93 | 812 | 0.971 | 5461 |  |  |  |  |  |
| Stomach | 68 | 812 | 0.978 | 163 |  |  |  |  |  |
| Pan-cancer | 1004 | 812 | 0.926 | 1 |  |  |  |  |  |

### Comparison of ROC-based biomarker discovery to commonly used feature selection methods

Reduction of the feature space by eliminating non-informative and enriching informative measurands in a data set is typically the first step of biomarker discovery. Methods commonly used to this end include variants of regression analysis, the machine learning technique Random Forest, the linear model of principal component analysis (PCA), and association analysis by Spearman rank correlation. However, the impact of the choice of method to select the most promising biomarker candidates for validation is unknown. To understand whether these methods enrich the same features as the ROC, we determined their ability to find biomarkers with sufficient ROC AUC for use in screening applications in data from the CancerSEEK discovery study. We did an exhaustive search by computing the ROC AUC values, with ∼3.9×10^19^ ROC points for all composite biomarkers of four measurands in the dataset. We then used feature selection methods to reduce the number of four-measurand biomarkers in the original datasets by a factor of 100. We set the ROC AUC of the FIT test used for colorectal cancer screening as the divider between biomarkers of interest and those not of interest. Comparing the biomarkers selected by each method to the ROC analysis, the best feature selection method retained only 15% of biomarkers equal to or better than FIT when considering the AUC value, all while also selecting biomarkers with lower ROC AUC than FIT (Fig. 7A). When reduced by factors of 1, 000 and 10, 000, the fraction of biomarkers of interest included by the best performing non-ROC feature selection method decreased to <3% and <1%, respectively (Fig. 7B).

**Fig. 7.**
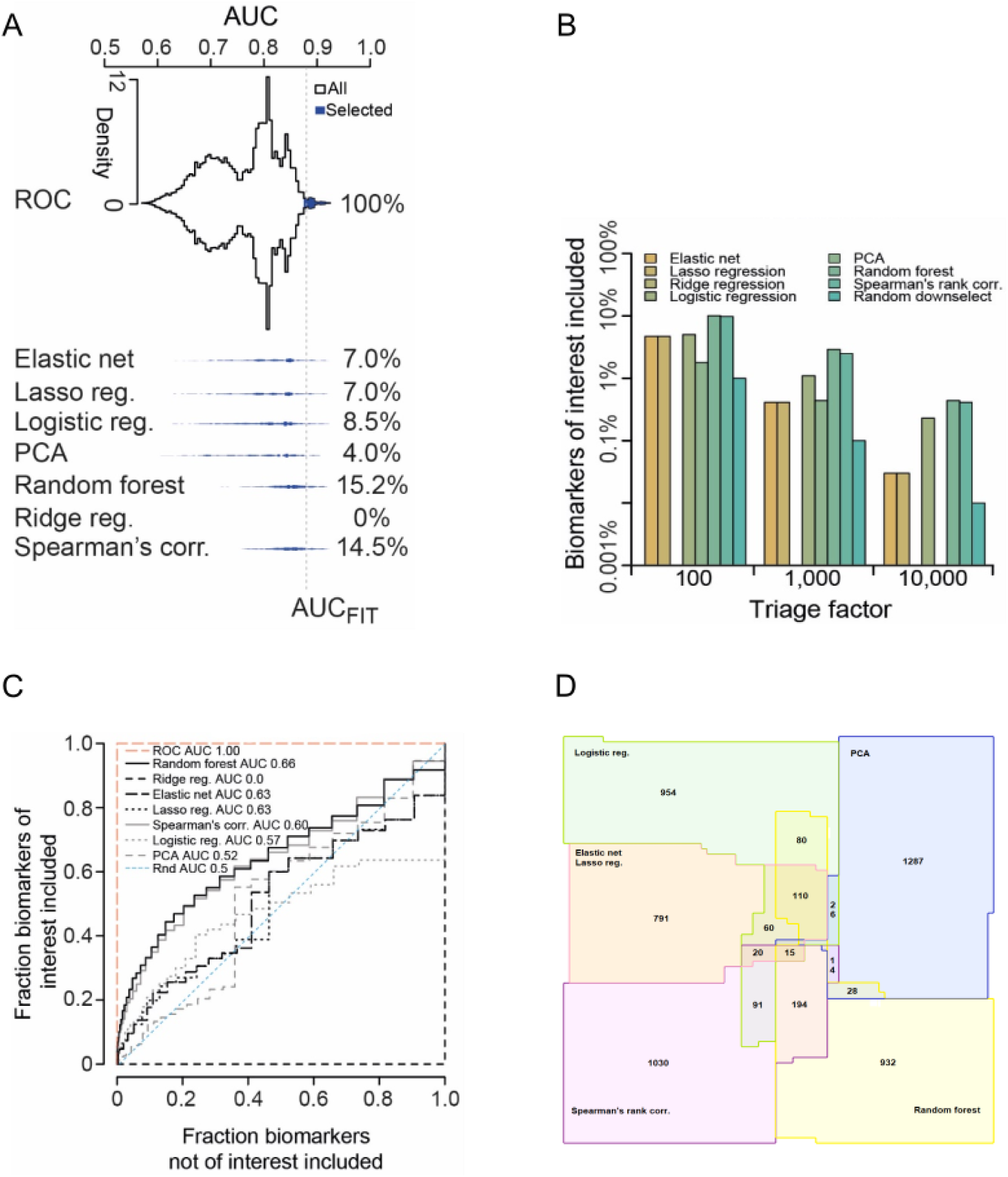
Feature selection methods commonly used in biomarker discovery perform poorly as surrogates for ROC AUC. **A.** Composite biomarkers were formed by combining the measurands in CancerSEEK data in combinations of 4. Triage by statistical methods commonly used in biomarker discovery studies was compared to informed downselect based on ROC AUC values in the ability to select biomarkers of equal or better performance than a predicate device (FIT, ROC AUC 0.88). Distribution of ROC AUC values of composite biomarkers (violins) and the putative biomarkers selected by each method at a triage factor of 100 (blue). Values indicate the fraction of selected biomarkers with ROC AUC ≥ FIT (hatched line). **B.** Fraction of qualitative biomarkers (ROC AUC ≥ 0.88) included when decimating the total number of biomarkers by different feature selection methods and magnitudes. For example, triaging data set by a factor of 1, 000 using Random Forest yielded ∼5% of the biomarkers of interest as opposed to the list obtained when selected by the ROC AUC. **C.** ROC plots showing abilities of triage methods to include biomarkers of interest (ROC AUC ≥ 0.88) while excluding those not of interest (AUC < 0.88), in CancerSEEK data. Each step effectively represents an additional included measurand through a more liberally applied triage. For reference, the random downselect (Rnd) has AUC 0.5 while the informed downselect (ROC) has AUC 1. **D.** Limited selection overlap by triage methods commonly used in biomarker discovery studies. The statistical methods indicated were applied to all putative pan-cancer composite biomarkers formed by all possible combinations of 4 measurands from CancerSEEK data at a triage factor of 100. The overlap of selected biomarkers was visualized in an area representative Venn diagram. ROC, Receiver-Operator Characteristic; AUC, Area Under Curve; FIT, Fecal Immunochemical Test; PCA, Principal Component Analysis.

Evaluating how the estimated sensitivity and specificity of composite biomarkers selected by each feature selection method compared to those selected by our ROC AUC-based computation, we found that these methods all had moderate to low ability to select biomarkers with ROC properties suitable for regulatory approval processes for follow-up analyses (Fig 7C). Of the seven methods evaluated, Random Forest performed best, followed by Spearman’s rank correlation coefficient (AUC 0.61-0.65) (Fig. 7C). The other methods had AUC < 0.6 and occasionally < 0.5, meaning that they had worse ability of selecting biomarkers of interest than chance alone. Lasso regression, ridge regression and elastic net are closely related statistical methods, distinguished only by a parameterized penalty factor. In the CancerSEEK dataset, lasso regression and elastic net performed on par with random down select (AUC 0.51) and ridge regression failed.

When intersecting the sets of biomarkers identified by the different feature selection methods, no biomarker was selected by all, and the best pairwise overlap between any two methods was 17% (Fig. 7D). Sets of measurands may exhibit interaction effects, i.e. joint patterns that are different from the sum of their individual patterns. Here, PCA, random forest, and Spearman’s correlation coefficient performed similarly in the presence of interaction effects as in the absence, whereas elastic net, lasso regression and logistic regression performed significantly worse in the presence of interaction effects (Suppl. Fig. 3). Taken together, none of the seven frequently used feature selection methods performed well as a surrogate for the ROC AUC, highlighting the need for new approaches to biomarker discovery.

## Methods

### Ethical approval

The study was approved by the Swedish Ethical Review Authority (EPM dnr 2019-00222) with amendments. The research was in line with informed consents obtained from patients included in U-CAN (EPN Uppsala 2010-198 with amendments) and research participants in EpiHealth.

### Datasets

The CancerSeek study was used as a source of plasma biomarker data, generated by measuring 39 proteins in 1, 005 cases across 8 tumor types and 812 controls (31). In case of sex-specific cancers (e.g. ovarian cancer), we chose to only include healthy controls of the relevant sex.

The Swedish dataset consists of the levels of 165 proteins in plasma samples collected at diagnosis, prior to any treatment, from 268 colorectal cancer, 265 lung cancer, and 167 ovarian cancer patients with tumor stages I–IV. These samples were obtained from the prospective, population-based U-CAN cancer biobank. The controls consisted of plasma samples from 697 individuals in the EpiHealth cohort, ensuring they were within the same age range and residing in the same region as the cancer patients. To maintain relevance for cancer screening, cases and controls were aged 45–75 years for colorectal and lung cancers, and 40–80 years for ovarian cancer. For ovarian cancer, we chose to only include female healthy controls (Åkerrén Ögren et al, submitted).

### Generation of composite biomarkers from unitary constituents

Classification composite biomarkers (CCBs) are a natural extension of the unitary biomarker with a cut-off; applying one cut-off each to two or more constituent biomarkers. Using two constituent biomarkers, CCBs are discussed by Prakash et al. [25] and implemented in IVD devices such as Banyan BTI and Abbott i-STAT TBI [26]. We have extended CCBs to allow for arbitrary numbers of constituent biomarkers and developed computational algorithms for efficiently computing ROC AUC values for such composite biomarkers when the constituent biomarkers are either over or under their respective cut-offs under logical rules ALL (all constituents) or ANY (any of the constituents).

When the number of constituent biomarkers increase, for instance from 3 to 4, the number of composite biomarkers grows rapidly, following the binomial coefficient *n choose k*, which determines the number possible combinations of constituents. For each combination, multiple candidate biomarkers are generated based on threshold conditions (over or under a threshold) and by the application of the logical rules ALL or ANY. This effectively multiples the number of combinations with *2^k+1^*, where k is the number of constituents in a CCB, to account for all putative biomarkers. Additionally, computing the ROC AUC for each CCB increases in workload since there are more cut-offs to evaluate. All these factors contribute significantly to the considerable computational workloads. However, when juxtaposed with the cost of sample procurement and data production, the computational cost is typically non-dominant and is projected to decline gradually since computational resources historically have become more affordable over time.

An algorithm for computing ROC AUC value for CCBs through data may compute True Positive Counts (TPCs) and False Positive Counts (FPCs) for every n-tuple of cut-offs. If data are sorted per one of the constituent biomarkers, counts for said constituent biomarker can be obtained through integer increment, which is a computationally efficient operation, while the other constituent biomarkers are evaluated for each data point via comparison. Other computational efficiencies are obtainable [32].

### Statistical simulations

We used Monte Carlo (MC) simulations in various aspects of the study. For each MC simulation a chosen number of cases and controls was drawn at random from a bi-normal distribution with defined form and particular position of the modes in the interval [0, 1]. Following individual simulations ranging in number from 1×10^8^ to 1×10^10^, we obtained the density distribution in each simulated scenario. Thus, we were able to numerically compute several probability-related functions characterizing the underlying distribution, such as probability density function, cumulative distribution function and most importantly inverse cumulative distribution function (quantile function). The quantile function was used to estimate the statistical significance of each putative biomarker or in other cases the statistical power of the simulated scenario.

### Statistical power and sample allocation optimisations

The statistical power was estimated as a function of study size and sample allocation to training and validation sets using MC simulations. Sample sizes ranged from 50 to 400 in increments of 50. The primary evaluation criterion was the number of putative biomarkers advancing to validation, assessed through 1×10^8^ MC simulations for each sample size or allocation strategy. By simulating different case-to-control ratios, we determined that an optimal ratio of 1:1 yielded the best performance and was used for subsequent optimizations. We simulated the number of biomarkers on par with ColoGuard (AUC 0.93) that had to exist in the data to discover and validate at least one of them given different study sizes or various sample allocations. While operating under the conservative scenario, when only a single true ColoGuard-class biomarker exists in the data, we estimated the targeted statistical power across the range of biomarkers advancing to the validation step.

For scenarios with condition-specific controls, we estimated statistical power in comparisons of cases versus matched controls and cases versus pooled controls. When control cohorts were pooled, multiple hypothesis testing was accounted for by adjusting the statistical power with a Bonferroni correction.

### Optimization of study design by using a predicate device

Statistical power is the probability of observing a putative effect being statistically different vis-à-vis the null-hypothesis effect. Choosing as null-hypothesis the performance of a relevant predicate *in vitro* device aligns well with regulatory practice. For example, for colorectal cancer (CRC) the IVD device Epi proColon (AUC 0.82) [33], was at time of writing the highest performing regulatory approved blood test for detection of CRC and cancer in general.

The probability distributions for the ROC AUC value, under various sample sizes (from 50 to 400) for cases and controls, were obtained through the method described previously [16]. The continuous AUC-value probability distribution and the respective quantile function were computed through 10^10^ Monte Carlo simulations, while 60 AUC probabilities were calculated exactly, and geometric interpolation was performed to the 99.99^th^ percentile. For robustness, several null-hypotheses were investigated; performance on par with ROC AUC values 0.76, 0.82 and 0.88, the latter corresponding to CRC stool test FIT. The putative effects investigated were performance on par with ROC AUC values 0.82 (for null-hypothesis ROC AUC 0.76), 0.88 (for null-hypotheses ROC AUC 0.76 and 0.82), and 0.93, which corresponds to CRC stool test ColoGuard (for null-hypotheses ROC AUC 0.88 and 0.82) [28]

The probability of success in the training step, i.e. successfully advancing the putative biomarker to the validation step, amounts to the probability of observing the putative biomarker ROC AUC value higher than the k^th^ highest observed ROC AUC value of the null-hypotheses effects, where *k* is the number of biomarkers that are advanced to the validation step. The k^th^ highest observed ROC AUC value is efficiently obtained using order statistics, through sup{x: F(x) ≤ U} where F is the cumulative distribution function of the ROC AUC value of the null-hypothesis effect and U an observation from the Beta(n+1-k, k) distribution where n is the total number of biomarkers included in the training step. By the principle of most conservative assumption, all biomarkers in the training step that are not a putative biomarker are on par with the null-hypothesis effect biomarker. If the sample size in the training step is zero, then the training step reduces to a random downselect and consequently the probability of successfully advancing the putative biomarker equals *k/n*, where k and n are as above. In particular, if *k=n*, i.e. all biomarkers in the training step is advanced to the validation step, then *k/n=1*. The probability of success in the training step was estimated using 10, 000 generated observations of U through the function rbeta in the stats package of R (version 3.3.2).

The probability of success in the validation step, i.e. observing a putative biomarker ROC AUC value that is higher than the critical value of the one-sided null-hypothesis ROC AUC value at statistical significance level 99% when Bonferroni-corrected for the number of biomarkers advanced to the validation step, is obtained through 1-G(sup{x: F(x) ≤ 1-0.01/k}) where G is the cumulative distribution function of the putative biomarker ROC AUC value and F and k are defined before. Finally, because distinct data are used for the training and validation steps, the two are statistically independent, and consequently the statistical power equals the product of the probabilities of success in the training and validation steps.

### Biomarker discovery in CancerSeek data and validation in a Swedish cohort

We computed and evaluated ROC AUC of all composite pan-cancer and tumor type specific biomarkers composed of four measurands, for a total (39 choose 4) = 82, 251 combinations, resulting in 82, 251 × 9 (groups) × 32 (rules) = 23, 688, 288 putative composite biomarkers. For each classification scenario, an exhaustive search was conducted to evaluate all possible cutoff values and optimize for the best possible ROC curve. This involved maximizing the true positive rate for every false positive rate to achieve the highest possible AUC. We then performed statistical hypothesis testing to determine whether the biomarker differentiated cases from controls more effectively than the benchmark biomarker Epi proColon, which is an FDA-approved diagnostic blood tests for colorectal cancer. The comparison was conducted at a 99% significance level, following the same threshold and rules used in the analysis of cancers versus controls. This process was repeated for each tumor type. We applied Bonferroni correction to adjust p-values, accounting for all computed combinations of the respective cancer type. For external validation of protein biomarkers in the Swedish cohort, we calculated the ROC AUC for the combinations of all statistically significant biomarkers from CancerSeek dataset, following the described rule set and employing the same methodology. During the composite biomarker computation, cutoff values were optimized independently in each of the datasets. This approach ensured that while the same protein combinations and regulatory patterns (up/down-regulation) were compared, absolute cutoff values were not, mitigating baseline differences caused by variations in experimental protocols or sample handling.

### Benchmarking commonly used feature selection methods to exhaustive search by ROC

A good feature selection method will select a large fraction of biomarkers of interest while selecting a small fraction of biomarkers not of interest, and we therefore used the estimated sensitivity and specificity of the composite biomarkers selected by each triage method to evaluate the methods like diagnostic tests by ROC analysis using the exhaustive search as ground truth. The ROC AUC analysis was compared to seven feature selection methods: Spearman’s rank correlation, Logistic regression, Principal component analysis, Elastic net, Lasso regression, Ridge regression and Random Forest. Each method was applied to the CancerSEEK data as a R-function with options and parameters described in Suppl. Table 2.

## Discussion

When considering the different statistical methods used in biomarker discovery, it is not known whether any or all will select the best candidate biomarkers considering the ROC based performance requirements for clinical use. Here, the top performing such method, Random Forest, selected on average only 2.2% of the biomarkers of interest when triaging by a factor of 1, 000, and Spearman’s rank correlation performed second best (1.7%). Inability to handle interaction effects contributed to the subpar performance of elastic net, lasso and ridge regression, but not to that of the other triage methods. No triage method performed significantly better when using quantitative data relative to ranked data, which demonstrates that none of them can exploit the quantitative information in the data to accurately predict the ROC AUC values of composite biomarkers. Typically, these different methods are meritoriously used in epidemiological studies to find differences in averages between groups and identifying risk factors but they may be ineffective as surrogates for ROC AUC, the widely endorsed diagnostic accuracy figure of merit [10]. Given the poor performance of the evaluated triage methods and the limited overlap between the sets of biomarkers selected by each one of them, it is warranted to include their use among the known pitfalls of biomarker development To increase the likelihood of discovering biomarkers fulfilling regulatory criteria, we instead developed an approach based solely on the ROC statistic used by regulatory bodies for assessing diagnostic accuracy of IVD tests or diagnostic biomarkers.

We here propose a biomarker discovery study design conformant with the regulatory approval process for a diagnostic test. To help advance the translation of biomarkers, we conceived a framework for early discovery studies implementing best practices and experiences from the literature to mitigate all known shortcomings not related to pre-analytical properties of the samples or the selection of study subjects. This study design is comprehensive from formulation of biomarker study objectives through data analysis, and represents an attempt at Quality by Design, a phrase commonly used vis-à-vis preemptive measures employed to minimize bias. The clinical trial framework is built on time-tested and fundamentally sound principles, and an additional synergistic benefit is that designing the study to regulatory standards facilitates prospective development of a putatively discovered biomarker into a regulatory approved IVD device. As demonstrating non-inferior diagnostic accuracy of a new biomarker test to an already approved predicate device is fundamental to the regulatory approval process, it is reasonable to benchmark to such state-of-art devices already during early discovery studies. The proposed study design framework conforms to regulatory requirements as it uses ROC AUC exclusively as Fig. of merit for diagnostic accuracy, an approach that has been proposed [25, 28] but not previously demonstrated.

From our simulations, it emerged that (i) 300 cases and 300 controls was optimal for having >95% statistical strength of discovering one ColoGuard-class effect in a background of Epi proColon-class biomarkers, (ii) the ratio of cases to controls, ideally 1:1, has limited impact on statistical power in the range 1:2-2:1, (iii) statistical strength was best when all data was allocated to the validation step with hypothesis testing under Bonferroni correction, (iv) while adding controls will always increase statistical power, pooling of control groups beyond an optimum will entail loss of statistical power from multiple hypothesis testing.

Contemporary convention is to design biomarker studies such that data is divided into a training and validation step, where a subset of biomarkers are identified in the training set and subjected to validation in a validation set. The results presented here are clearly at odds with this convention since the implication in all simulated scenarios is that it is optimal to bypass the training step and advance all composite biomarkers to the validation step. Moreover, it can be argued that partitioning data produced under the same conditions and analyzing a fraction of the data for validation purposes is less stringent than performing validation using external datasets. Because the total number of composite biomarkers is limited, it is possible to correct for testing of multiple hypotheses throughout the range of one up to all of the biomarkers, and we demonstrate conduct of detailed analysis of sample sizes with consistency of study endpoints under a minimum of assumptions. These results are particularly relevant for -omics based efforts, as the study design allows for validation of many biomarkers under Bonferroni correction.

To demonstrate the study design concept, we analyzed publicly available CancerSeek data. We use data obtained through Luminex xMap technology, which is a multiplex-implementation of sandwich ELISA, to find and validate composite biomarkers. Using publicly available data sets, the total numbers of samples and controls are fixed and are sub-optimal for some tumor types in the simulations performed here. It may also be noted that the original CancerSeek study did not include statistical hypothesis tests or p-values. Importantly, the pan-cancer biomarker identified here is a subset of the CancerSeek panel that had better ROC AUC performance than the combination suggested by Cohen et al [31].

As successful clinical development of a biomarker hinges on its ROC performance, it is advisable to rely on ROC analysis already in the early discovery phase. Using the tools provided here, the investigator can for the first time make informed decisions on key parameters of biomarker study design in favor of successful translation to IVD tests. While the approach is applicable to biomarker discovery in general, a limitation is that signature size scales with the computational intensity. On the other hand, its ability to find small signatures with high diagnostic performance has advantages as the robustness of an IVD test may suffer from each extra measurand incorporated. Finally, it is likely that useful biomarkers have been overlooked in otherwise appropriately performed studies which should motivate re-analysis using this novel framework.

## Data Availability

All data produced in the present study are available upon reasonable request to the authors

https://www.science.org/doi/suppl/10.1126/science.aar3247/suppl_file/aar3247_cohen_sm_tables-s1-s11.xlsx

## Acknowledgement

The study was funded by grants to T.S. from the Swedish Cancer Society (CAN 2018/772, 21 1719 Pj, 2024 3831 Pj), the flagship project grant from Sjöberg Foundation (2024-11-06: 18, 2) and Lena Wäppling’s foundation, and from the Swedish Cancer Society to J.Å.Ö. (CAN 21-0430-PT). We are grateful to U-CAN for access to plasma samples from Uppsala Biobank with support from the Swedish Government (SRA grant CancerUU), and EpiHealth for plasma samples collected with support from the Swedish Government through the Swedish Research Council

---

**Suppl. Fig. 1.**
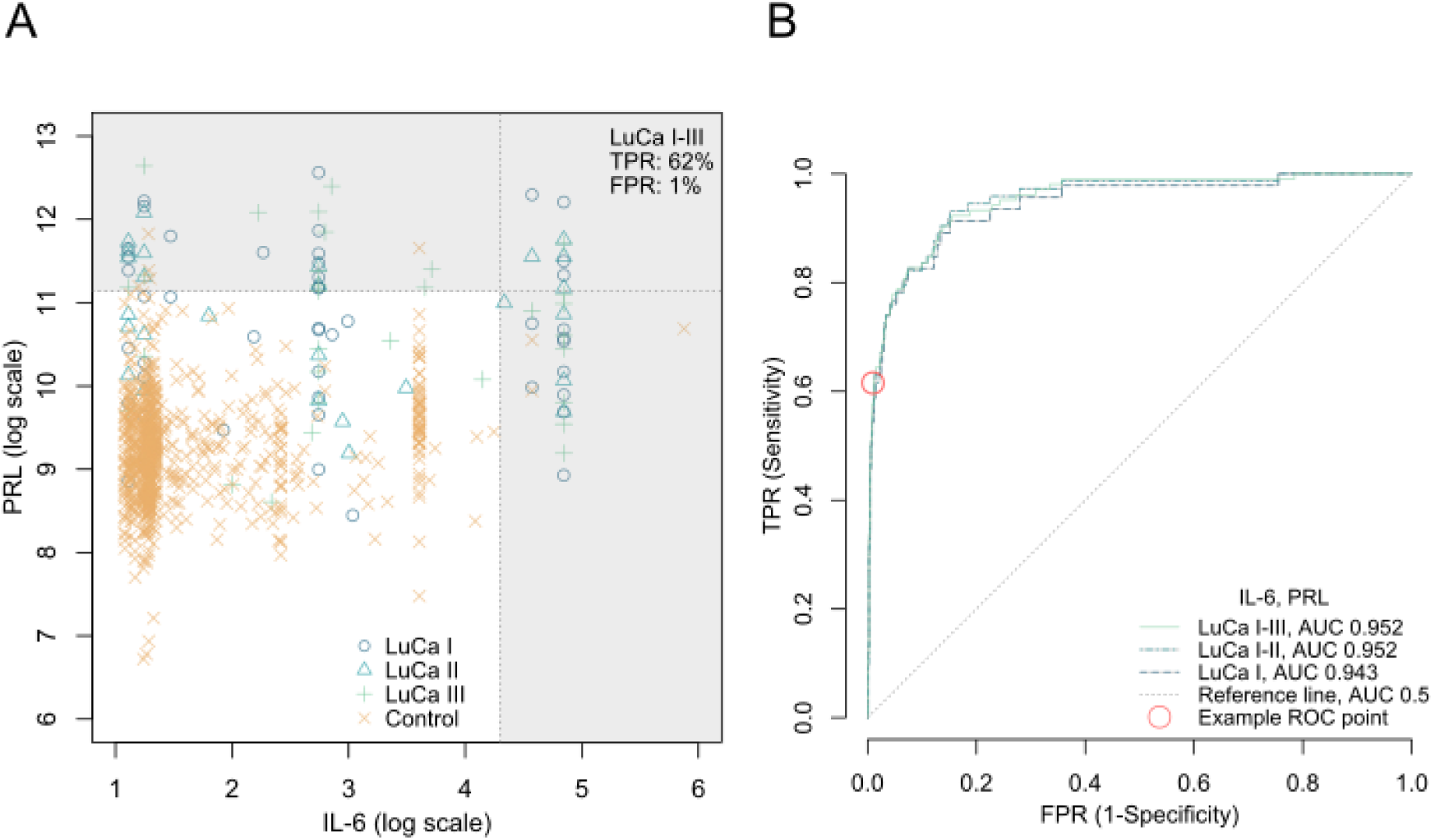
Relationship between classification region and ROC curve for a composite biomarker of 2 measurands. **A**. Scatter plot of PRL and IL-6 levels measured in LuCa patients and healthy controls in CancerSeek. Grey area, positive classification region where PRL exceeds its cutoff level or IL-6 exceeds its cutoff level. TPR and FPR constitute a ROC point for the composite biomarker IL-6 and PRL. **B**. By exploring all cutoffs of the composite biomarker, a ROC curve with AUC 0.952 for LuCa stage I-III is obtained. Red circle, ROC point of classification region shown in **A**.

**Suppl. Fig. 2.**
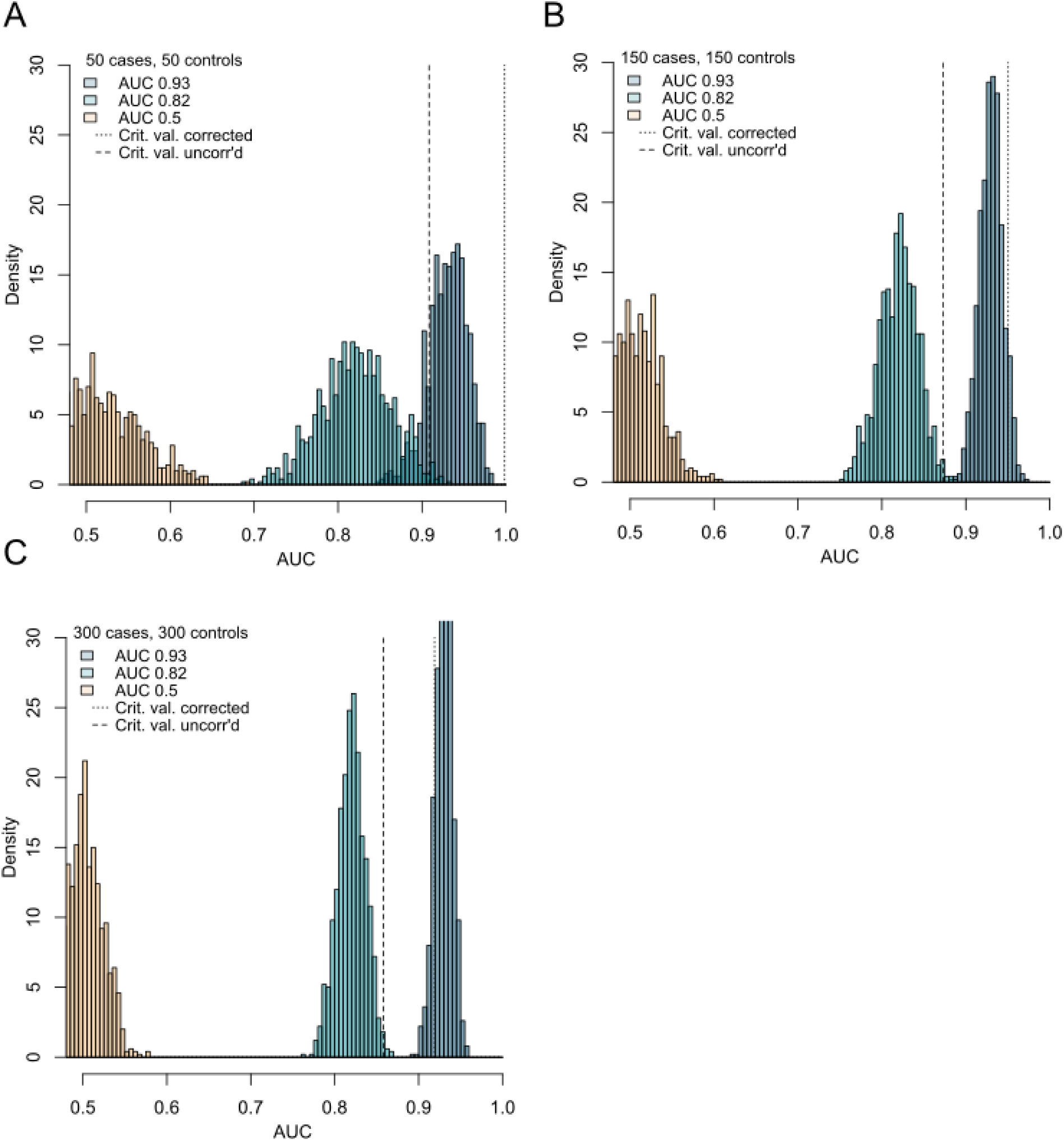
Statistical strength to demonstrate superiority of a biomarker on par with ColoGuard to a biomarker on par with Epi proColon under a range of sample sizes. Monte Carlo simulated ROC AUC values of biomarkers on par with Epi proColon (ROC AUC 0.82) and ColoGuard (ROC AUC 0.93) under different sample sizes. Critical value, observed AUC value greater than a biomarker on par with Epi proColon with 99% significance. Corrected, critical value after Bonferroni correction for multiple hypothesis testing. Fraction of blue bars (AUC 0.93) with AUC greater than the corrected critical value corresponds to the statistical power. **A.** 50 cases and 50 controls (0% statistical power). **B.** 150 cases and 150 controls (20% statistical power). **C.** 300 cases and 300 controls (96% statistical power).

**Suppl. Fig. 3.**
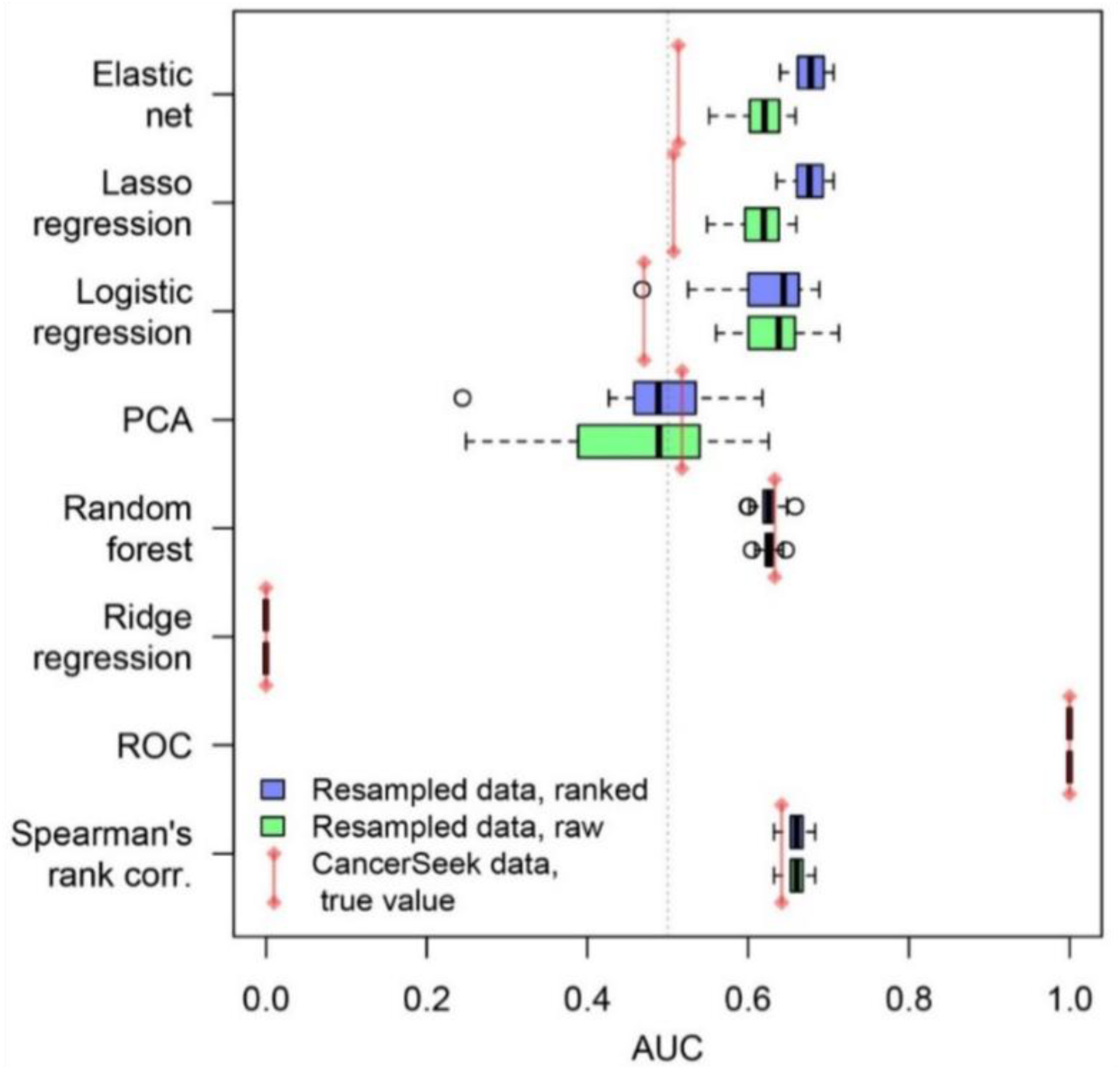
Performance of feature selection methods when interaction effects are eliminated by resampling of data under statistical independence. Box plots of ROC AUC computed for all combinations of four measurands from raw or ranked CancerSeek data resampled 25 times under statistical independence. If a triage method, such as e.g. logistic regression, performs equally well when using the ranks relative to using quantitative data, it follows that the model fitted is inept at executing its core purpose, which is to accept quantitative data as input and utilize them to produce outcome predictions of relatively high accuracy. Red bars, AUC performance with non-resampled CancerSeek data. Boxes, first and third quartiles. Whiskers, min and max values. Circles, outliers.

**Suppl. Table 2.** Methods and parameters used to assess triage methods. The parameters of each regression analysis method or machine learning technique evaluated in the paper are listed, together with the respective R-packages used in the study.

| **Triage method** | **Package, function (R version 3.4.3)** | **Non-default parameters** | **Variable forward selection method** |
| --- | --- | --- | --- |
| Spearman's rank correlation | *stats, cor* | *method="spearman"* | Greatest Spearman's rank correlation relative to binary outcome |
| Logistic regression | *stats, glm* | *family=binomial(link="logit")* | Least sum of squared model residuals |
| Principal component analysis | *stats, prcomp* | (none) | Least model standard deviation |
| Elastic net | *glmnet* (v. 2.0-16), *glmnet* | *family="binomial", alpha=0.9* | Sequentially increasing the parameter *pmax*, and identifying indices of the non-zero regression coefficients |
| Lasso regression | *glmnet, glmnet* | *family="binomial", alpha=1* | (see above) |
| Ridge regression | *glmnet, glmnet* | *family="binomial", alpha=0* | (see above) |
| Random forest | *caret* (v. 6.0-81), *trainControl*<br>*caret, train* | *method="repeatedcv", number=10, repeats=3, search="random"*<br>*method="rf", metric="Accuracy", tuneLength=15* | As per function *varImp* |

## Notes

### Competing Interest Statement

The authors have declared no competing interest.

### Funding Statement

The study was funded by grants to T.S. from the Swedish Cancer Society (CAN 2018/772, 21 1719 Pj, 2024 3831 Pj), a Flagship project grant from the Sjöberg Foundation (2024-11-06: 18,2) and Lena Wäpplings foundation, and from the Swedish Cancer Society to J.Å.Ö. (CAN 21-0430-PT). We are grateful to U-CAN for access to plasma samples from Uppsala Biobank with support from the Swedish Government (SRA grant CancerUU), and EpiHealth for plasma samples collected with support from the Swedish Government through the Swedish Research Council.

### Author Declarations

The analyses of the plasma samples were approved by the Swedish Ethical Review Authority (EPM 2019-00222).

